# Intervention Mapping: A Multi-stage investigation to Control Respiratory Tract Infections at Primary Level

**DOI:** 10.1101/2025.02.20.25322323

**Authors:** Shahzad Mahmood, Nauman Ali Chaudary, Mariam Nawaz, Mohsin Nazir, Muhammad Shahid Mahmood

## Abstract

Respiratory Tract Infections are the major health hazard to humanity, nowadays. This research aims at the development and implementation of evidence-based intervention at primary healthcare level, in order to control Respiratory Tract Infections, called Intervention Mapping(IM). It comprises multiple phases, focussing on specific needs of the target population and ensuring that intervention should be based on behavioral and systematic change. The study has a mixed design, quantitative and qualitative approach, involving multiple stakeholders, community’s participation and healthcare worker’s collaboration, to control infectious diseases at primary healthcare level and thereby to reduce the burden of diseases.

The study comprises three phases including a pre-intervention phase in which disease load was assessed and the need of particular intervention was analyzed. A multi-targeted public health intervention, incorporating health education, environmental modification, and policy changes, was designed and applied after community analysis. This phase is preceded by the insight development phase which showcased the vitality of community based intervention. The infectivity of respiratory tract infections is studied under two categories. The gender based segregation provided an insight on the spread of respiratory tract infections in males and females, and a subsequent reduction in disease load among both the genders. The second form of study is focussed of age based separation and disease load. The effectiveness of the research can be analyzed by a remarkable decrease in the respiratory diseases in a community.

According to the study’s findings, epidemiologic burden of respiratory infectious diseases can be trimmed by community strengthening activities including population sensitization, community health initiatives, targeting at-risk population, and engaging key actors of the area. This translational research provides a policy recommendation for health promotion, disease prevention, and intervention mapping in a resource-constrained environment. However, the pathway stipulated by the study revolves around shifting from a curative model of healthcare to the preventive model at primary health level.

## INTRODUCTION

Two centuries ago after Louis Pasteur promulgated that all infectious diseases could be prevented and that diseases develop after the contamination by invisible microorganisms (Germ Theory of disease 1861), a question looms: If people are still suffering from infectious diseases after knowing the root cause, what kind of progress have humans made? Around 27% of the world population is suffering from the diseases caused by the spread of infections (Michaud 2009), and when their propagation rate increases, an epidemic becomes inevitable. Therefore, there is a pressing need to devise mechanisms to contain, if not mitigate, the dissemination of infection at primary level through research and development.

### Background of the Study

The science and art of public health has been using intervention mapping for years and there are thousands of articles published in this domain. Intervention mapping(IM) is a simple technique used to draw a practical community health approach by connecting it to a theoretical framework of evidence-based public health promotion programs. It is an amalgamation of theory and practice with a sequential roadmap for intervention development, implementation and evaluation. Community participation holds a key significance in the research because it guides about the need of particular intervention, from personal to community level, and the mode of intervention and evaluation.(Fernandez 2019)

Over the past two decades the process of IM has gone through significant improvement and a sophisticated theoretical groundwork has laid the foundation of public health which provided a handbook for practical steps to control infectious diseases. However, each study is faced with a myriad of limitations. The research to stipulate preventive measures against a pandemic, for instance, was based on a health belief model and was dealing with preventive measures at individual level. It lacks in interpersonal and community level analysis which are the cornerstones for disease prevention at a community level.(Tong 2020)

Similarly, another study entailing an IM program to control respiratory tract infection addresses the importance of health education ,but undermines the impact of social and environmental factors while designing an intervention.(Heshmati 2022) Therefore, the gaps in philosophical approach led to a realistic increase in infectious diseases specifical: the respiratory tract infections. This is evident from Covid pandemic 2019, the Swine Flu pandemic 2009, Ebola virus disease in West Africa 2013, Zika virus and countless epidemics. (Baker 2021) Pakistan is also affected by these global trends as reported by the World Health Organization that the country is among the top five states with tuberculosis endemicity. (WHO 2022) The intensity of the issue necessitates the assumption that there should be a study with strong philosophical foundations and can draw a clear map of interventions necessary to cope with the dissemination of infectious diseases.

### Statement of the Problem

In Rural areas of Pakistan, basic health units are functioning on curative model with a least focus on preventive measures and there is compelling need to focus on devising an evidence based preventive intervention rooted in Health Belief Model (HBM), Social Cognitive Theory (SCT), and Ecological Theory (ET). Despite implementing several strategies for health promotion at grassroot level, the prevalence of airways infections has remained alarmingly high. Consequently, high morbidity rates are echoing along with the burden of healthcare cost over families, community and the government.The root cause of this dilemma can be traced in inadequate health education at primary level, insufficient environmental sanitation, lack of community participation and paucity of translational research (Saqib 2021) at primary level. The study aims at fulfilling this knowledge gap by a three-phase intervention model.

### Objectives of the Study

The core objective of the study is to design a multiphase research methodology, IM, and then evaluate its effectiveness after its implementation at primary health level. However, the operational objectives include:

- To analyze the prevalence of respiratory tract infection in a rural area and segregation of the patients on the basis of age and gender
- To assess multiple factors involved in propagation of respiratory tract infection (RTI) at primary healthcare level
- To craft and evidence based intervention framework focussing on the control of the spread of infection
- To engage community and stakeholders for behavioral, systematic and environmental change that is vital to reduce the burden of disease
- To analyze the impact of intervention in terms of the change in prevalence of respiratory diseases
- To propose a potential and workable mechanism for disease prevention in the form of policy recommendations

### Research Questions

The viral research question is,

- What is the effectiveness of the multiphase intervention mapping(IM) approach in reducing the prevalence of respiratory diseases at primary healthcare level?

While answering this question many other curies require worth mentioning. These are:

- What is the prevalence rate of respiratory tract infections in the area under a basic health unit and how can this disease load be minimized?
- What factors are responsible for the propagation of infection there?
- How can community engagement be pivotal in hindering the spread of infection?
- Can a basic health unit become a precedent and present a model for the control of respiratory tract infections at national and global level?

### Research Hypothesis

The study is based on a directional hypothesis that is,

> “By Intervention Mapping, healthcare workers can reduce the prevalence of respiratory tract infectious diseases by 20% after two weeks of effective implementation of a community health program”.

### Significance of the Study

The undeniability of research’s prime importance is depicted by the strong theoretical foundations that are the bedrock for neo-innovation and future research. Intervention Mapping (IM) enriches the ideational framework for infection control particularly in resource-constrained settings where cost-effective mechanisms are direly needed for disease control. It provides an interdisciplinary platform of action for health promotion as,

**Fig 1.1:**
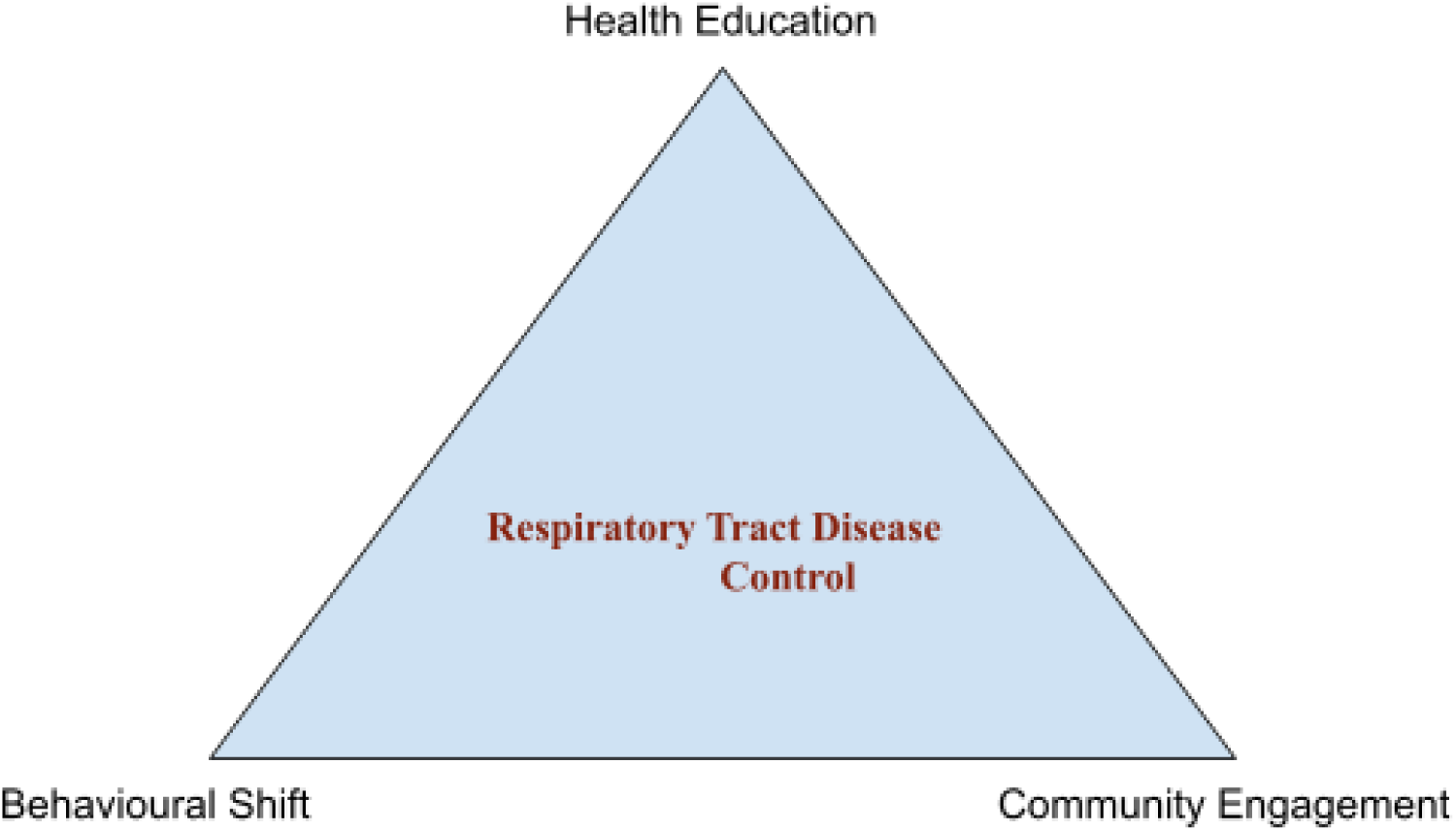

Besides providing a conceptual framework, the study has its distinctive practical significance and wide-range advantages of its implementation. It empowers rural communities to abate the threshold of microbial spread; thereby improving quality of life and reducing morbidity and mortality. It informs healthcare workers to become proactive in mitigation of infection rather than being reactive to cure a disease. The strategic shift is necessary especially in the geographies under an economic crisis because a drop in disease frequency will reduce the capital expenditure over the curative health sector. Thus, the study is a ray of hope for the crumbling health sector in the form of actionable policy recommendations for regulators and strategic planners.

### Ethical Consideration

The research is conducted by giving a key importance to ethics and morality. No participant was forced to join the research and the will of people was central in decision making. The qualitative surveys conducted were aimed at knowing the general acceptance and behavior of people towards a program. Since, the program was for public interest so it needed to be as according to the desire of people and it was given significant weightage. The privacy of people was kept intact; their confidentiality was never breached; and no illegality was committed during the whole project. The consent of people was taken before data collection, interviews and in every phase of research a right to consent was primary before taking any step. The study was conducted with a neutral approach by keeping the researchers unbiased and no tampering was made in any sort of data.

## LITERATURE REVIEW

### Intervention Mapping

The evidence based approach to plan, implement and evaluate a health promotion program is termed as Intervention mapping. Community involvement, participation of key actors and localization of strategy are the key ingredients of the technique which encompasses six steps of health promotion. (Garba 2017)

These are,

**Fig 2.1:**
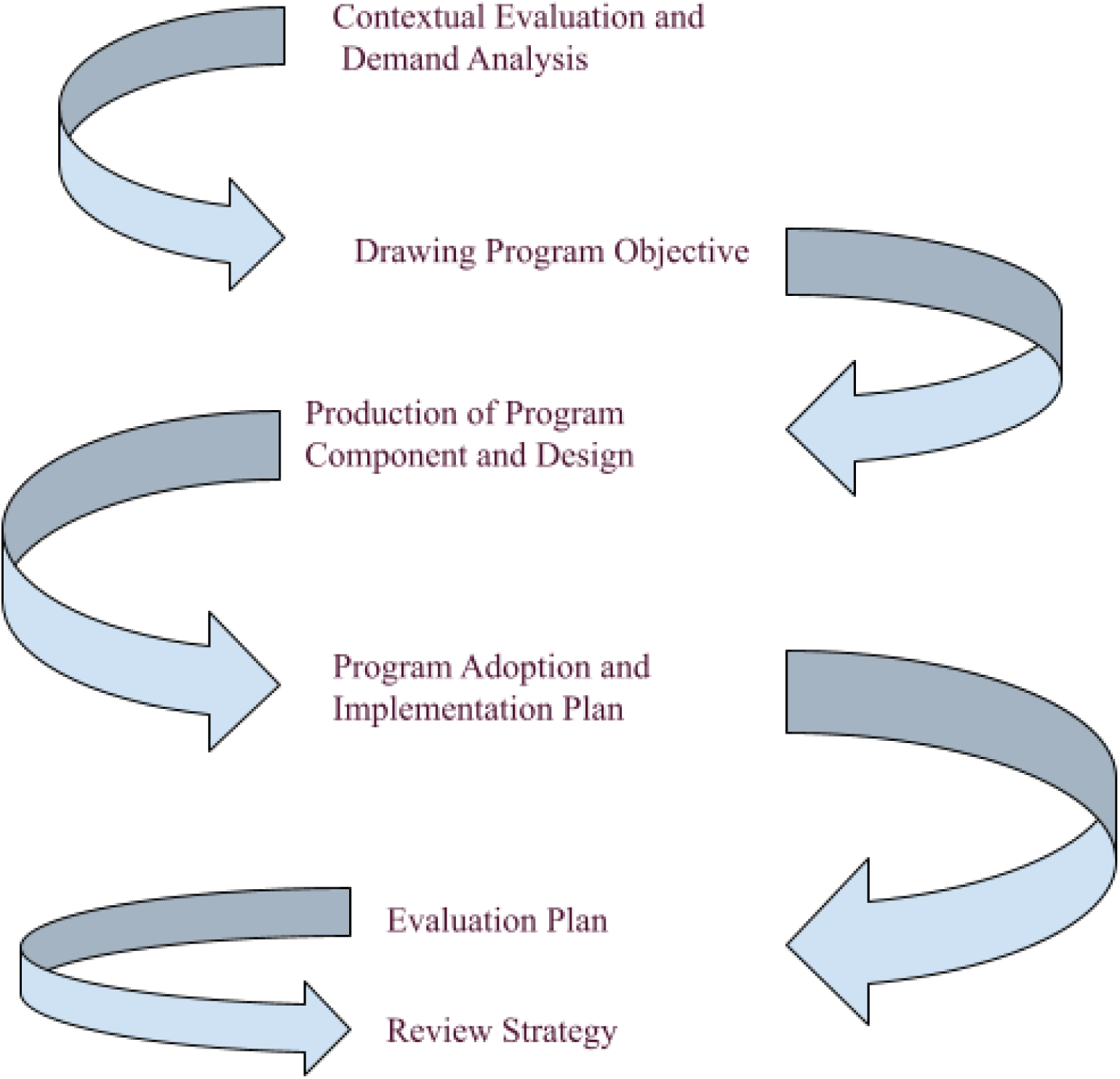

### Control of Respiratory Tract Infection

Personal hygiene plays a pivotal role in controlling respiratory infections. They are usually droplet infections caused by microbes spread through air. By sneezing and coughing the infected person spreads an infectious agent to a healthy one and thereby increases its prevalence by causing disease in the person. Pneumonia and Tuberculosis (Tb), as an illustration, propagates through droplet infection and Pakistan is an endemicity of Tb along with sporadic spikes of Pneumonia. The reason behind is inadequate health education in the population and the lack of awareness. People in rural areas do not pay any attention to their oral hygiene and, unintentionally, not only catch a disease but also play a role synonymous with the vector in the spread of infection. (Sjogren 2008)

The Upper Respiratory Tract infections are widespread in the countryside. Poor hand hygiene is the major cause of their dissemination. In rural areas, children are mostly affected by the URTIs which include both bacterial and viral infections like Influenza ,Rhinovirus and parainfluenza. A diseased child, after contaminating his hands with mucosal or serosal secretions of his nose, mouth and eyes, when touches a healthy child, he transmits the disease to the healthy child and the cycle continues till an incidental disease turns into an endemic and may subsequently leads to a pandemic, as has happened with H1N1 flu pandemic in 2009. (Leung 2023)

Besides epidemiology, the course of disease can also get worse. URTIs are, no doubt mostly, self limited, but can also yield serious complications such as Otitis Media, Bronchitis, Meningitis, Encephalitis, and Pneumonia. Infants and young children are the most susceptible to these morbidities because they do not have strong immunity to fight against the pathogenicity and virulence of the causing agent. Resultantly, mortality rates also increase with an increase in URTIs. (Sirota 2021) Apart from personal hygiene, environmental hygiene and URTIs also have a symbiotic relationship; that a healthy environment will make a disease attack less likely and vice versa.

Environmental hygiene includes, besides ecological sanitation, hygiene of the community and people surrounding a person called social hygiene. Poor sanitation, air pollution (bad air quality index) and particulate matters (PM) can accelerate the spread of disease. Particulate matter, for instance, can penetrate deeper in alveoli and can carry microbes with it. This can trigger alveolar damage and can produce a respiratory infection. (Khan 2024) Similarly, social hygiene is also of great importance. The community of smokers is seen to be more infected with respiratory diseases than the non-smokers. (Jiang 2024) It is not only because of carcinogens and irritants inhaled during smoking, but also due to cigarette sharing.

Sharing of contaminated material ,i.e cigarettes,vapes, utensils, stationery, and anything can be a source to disperse and cause viral illness. There arises a need to contain microbial spread through a target oriented approach also known as translation efforts in which an end goal to eradicate a disease is given the prime focus. (Olsen 2015) It may sound ironic but if smallpox can be rooted out from the globe and Polio can be in the last phase of its eradication, the respiratory diseases can also be minimized ,if not be wiped out, from the map of the world by hindering the spread of infection.

## STUDY DESIGN AND METHODOLOGY

### Research Design

This is a complex intervention design based on the model of intervention mapping at different ecological levels such as individual, family, group, and community. Each level is analyzed for its specific needs and intervention required. The whole study encompasses three phases consisting of six components.The first phase also known as exploratory analysis deals with three sub-level of intervention mapping. The first one is contextual evaluation and demand analysis that deals with an assessment of disease load and required intervention. The second sub-level is named as drawing program objective and the third one is production of program components and design.

The second phase also called Program Intervention is very crucial and it consists of only one component that is Program Adoption and Implementation Plan. After successful completion of this phase, the last one is titled as Evaluation and Impact Analysis Phase in which there are two sub levels designated as evaluation plan and review strategy.

### Study Focus and Sample Population

The study is conducted at a Basic Health Unit in Punjab, Pakistan. It is the primary pillar of primary healthcare in the state. It is a primordial presentation site of patients from nearly 10 villages and coverying 33432 people in a geographical zone of less developed area. The sample conduction site was a general OPD of 24/7 BHU and the intervention site was its field, covering 10 villages in its vicinity. Patients were segregated on the basis of their age and gender and they were asked questions about how they think they were infected and how they think they could have prevented this infection. This was the qualitative component of the study while the quantitative component deals with primary epidemiology of respiratory tract infections and the subsequent reduction in their disease load after an intervention.

### Timeline of the Research

The research was carried out in three and half months. The data collection for phase one was initiated on 15th of september, 2024 and carried for 45 days till 30 october, 2024. The Intervention phase had a span of 15 days and then the final phase continued from 15 November 2024 to 30 December. The timeline is ample illustrated in the table as,

**Table 3.1.**

| **Phases** | **Phase Name** | **Date** | **Timel**<br>**ine** |
| --- | --- | --- | --- |
| Phase 1 | Exploratory Analysis | 15 Sep, 2024 to<br>30 Oct, 2024 | 45<br>days |
| Phase 2 | Program Intervention | 1 Nov, 2024 to<br>15 Nov, 2024 | 15<br>days |
| Phase 3 | Evaluation & Impact<br>Analysis | 16 Nov, 2024 to<br>30 Dec, 2024 | 45<br>days |
| Final Report | Thesis Presentation | 10 Jan, 2024 | 10<br>days |

Total Duration: 3 Months and 25 days

### Exploratory Analysis (Phase 1)

The onset of the first phase marked the start of a new arena in the healthcare sector of the research area. It was commenced on 15th of September 2024 with an aim at analyzing the epidemiology of respiratory tract infectious diseases and to carve a way to reduce their prevalence. It further consisted of three components covering the two kinds of research paradigms such as both quantitative reviews and qualitative analysis. These are,

### Contextual Evaluation and Demand Analysis

This deals with analyzing the epidemiology of RTIs in different villages under the surveillance of the basic health unit. The quantitative sub-component of phase 1 focuses on collecting data about the prevalence of infectious diseases and their segregation according to age and gender. Data is collected from the general out-patient department of the BHU and the number of patients presented with a particular disease is recorded along with the name of their village or vicinity. This provided the researchers with an opportunity to draw the estimated distribution of particular diseases, their endemicity and infectivity.

The Respiratory Tract Infections reported from 15 Sep, 2024 to 30 Oct can be tabulated according to age and gender distribution as shown,

**Table 3.2.**
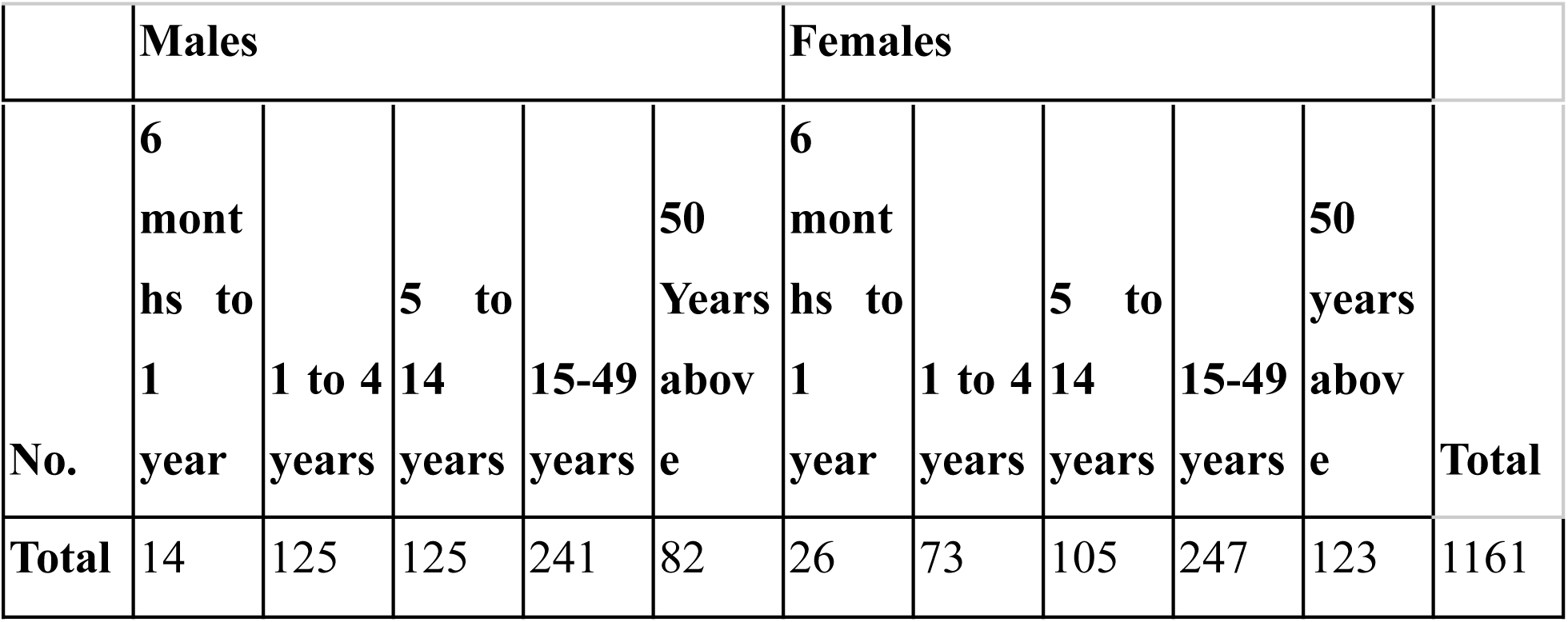
Age & Gender Distribution of RTIs(Before Intervention)

### Program Intervention (Phase 2)

As the heart is to the body, this phase is to research. The ideological roots of the phase lie in program adoption and implementation sub-component of Intervention mapping. A complex intervention designed after quantitative need assessment and qualitative surveys was applied on the population with an aim at minimizing the spread of infection through a public health remedy.This phase was spanned on 15 days from 01 Nov, 2024 to 15 Nov, 2024. The key interventions designed were,

### Intervention to Control Respiratory Tract Infections

Respiratory Tract Infections were most prevalent in the age group of 15 years to 50 years as 241 patients were reported in this age bracket during phase 1 of the research. The age bracket of 1 year to 4 years and 5 years to 14 years were also susceptible and 125 patients of each bracket were reported. Therefore, an intervention was drawn keeping a prime focus at these age groups. Qualitative surveys provided the researcher with a direction on what kind of public health program can effectively contain the spread of respiratory diseases. The program applied was,

#### Community Level Intervention

A basic health unit was dealing with 7 villages and some sub-rural attachments. Health Education and community sensitization were made a prime focus. To achieve these ends, seven community level organizations were formed, one in each village, known as RTI Leagues. Each was consisting of healthcare workers including a Medical Officer, Lady Health Supervisor, Lady Health Workers of that area, School Health & Nutrition Officer, and Sanitary Inspector along with key gate keepers of the community including Imam Masjid, village headman(lambardar), five males and five females (one from each street), and some notables as according to the designated village.

The role of RTI leagues was to provide health education in the community. An organizational constitution that provided the road map for action was drawn and all the activities were performed according to this Planning. Seminars were conducted in all villages by the RTI leagues. Home visits were paid so that people who could not attend the seminars can have a health education. After each prayer, imam masjid had delivered an address, as planned, to contain respiratory tract infections. LHW played a pivotal role in this program delivering the message of RTI league in the door to door campaign. The macro-level system was a mimicry of HIV control scheme in Africa in which text messages,using cell phones, were used to deliver the message of the use of condoms to prevent HIV.

Health Education was the bedrock of this health campaign. It included multiple components. Hand washing techniques are demonstrated in seminars and household campaigns. People were made aware of risk factors associated with the spread of infection and they were advised to wash hands frequently after encountering a risk factor. Personal hygiene was greatly stressed because almost all the respiratory infections are droplet infections that disseminate through coughing, sneezing, inhaling infected air, and using contaminated utensils.

Therefore, people are advised to use masks to cover their mouth when moving outside or even in the house when meeting with the other member of the family. Those who could not afford to buy a mask, since the basic health unit was situated in the most resource-constrained geography, are taught to use their turbans to cover their face, so that air borne transmission of respiratory infections can be hindered.Moreover, social distancing while greeting was practiced in demonstration and people were asked to follow this trend when greet.

#### Targeted Intervention

Besides community intervention, RTI league was also empowered to reach specific age groups in which disease is most prevalent and can be prevented by simple measures. For the targeted approach, people were touted according to the needs of their age-group. Children, for instance, were told to practice personal hygiene along with mask usage and their mothers were advised about the community level intervention with particular focus on their young ones. Mothers were told to avoid bottle feeding and to completely disinfect the bottle before feeding. They were told to practice chest physio of the child younger than one year.

Similarly, during the first phase, it was noted that the majority of the young and adults presented to the OPD with respiratory tract infection had a history of smoking and cigarette sharing. Therefore, during the intervention phase RTI league was also encouraged to target the people in the age bracket of 14 years to 49 years and educate them about the severity and the risk of infection associated with cigarette sharing. Besides mask usage and improving personal hygiene, the old people, those above 50 years of age, were advised, after demonstration of, respiratory exercises such as belly breathing and huff coughing and chest physiotherapy regularly.

### Evaluation & Impact Analysis Phase (Phase 3)

The final phase of the study consists of analysis of intervention impact, Impact evaluation, process evaluation and process feasibility. It encompasses the fifth and last sub-components of intervention mapping named evaluation plan and review strategy respectively. The basis of this phase is to evaluate the effectiveness of the intervention applied in phase 2.

### Intervention Impact Analysis

In order to determine the change in disease load after the intervention, the last phase started with the data collection about the number of patients reported with different diseases. The phase started from 15th September and lasted till 30th October 2024. The number of the patients reported in these 45 days were tabulated.

*The number of patients reported with respiratory tract infection were,*

**Table No. 3.3.**
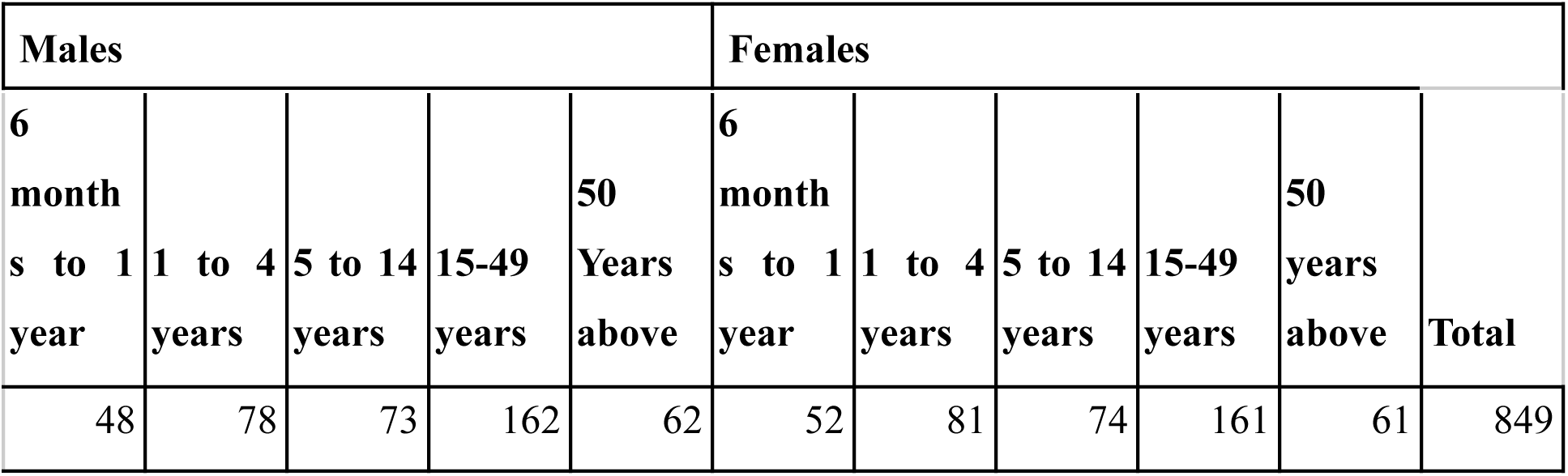
Disease Load after Intervention.

## Data Analysis & Interpretation

The data thus obtained by meticulous biostatistics in different phases of intervention is analyzed thoroughly.

### An Analysis of Pre & Post Intervention Respiratory Tract Infection

Respiratory Tract Infections were most prevalent in the geography of research, but after an intervention a considerable decrease in the number of these cases is seen

. As biostatistics show,

Decrease in the No. of Respiratory Infection Cases = No. of the Cases Reported in Phase 1-No. of the Cases Reported in Phase 3

=1161-849

=312

Percentage decrease in the No. of the Cases= (Actual Decrease/Total No. in Phase 1)100

=100(312/1161)

=26.873

*So, a 26.873% reduction in the cases of respiratory tract infections has been witnessed*.

**Graph 4.2.**
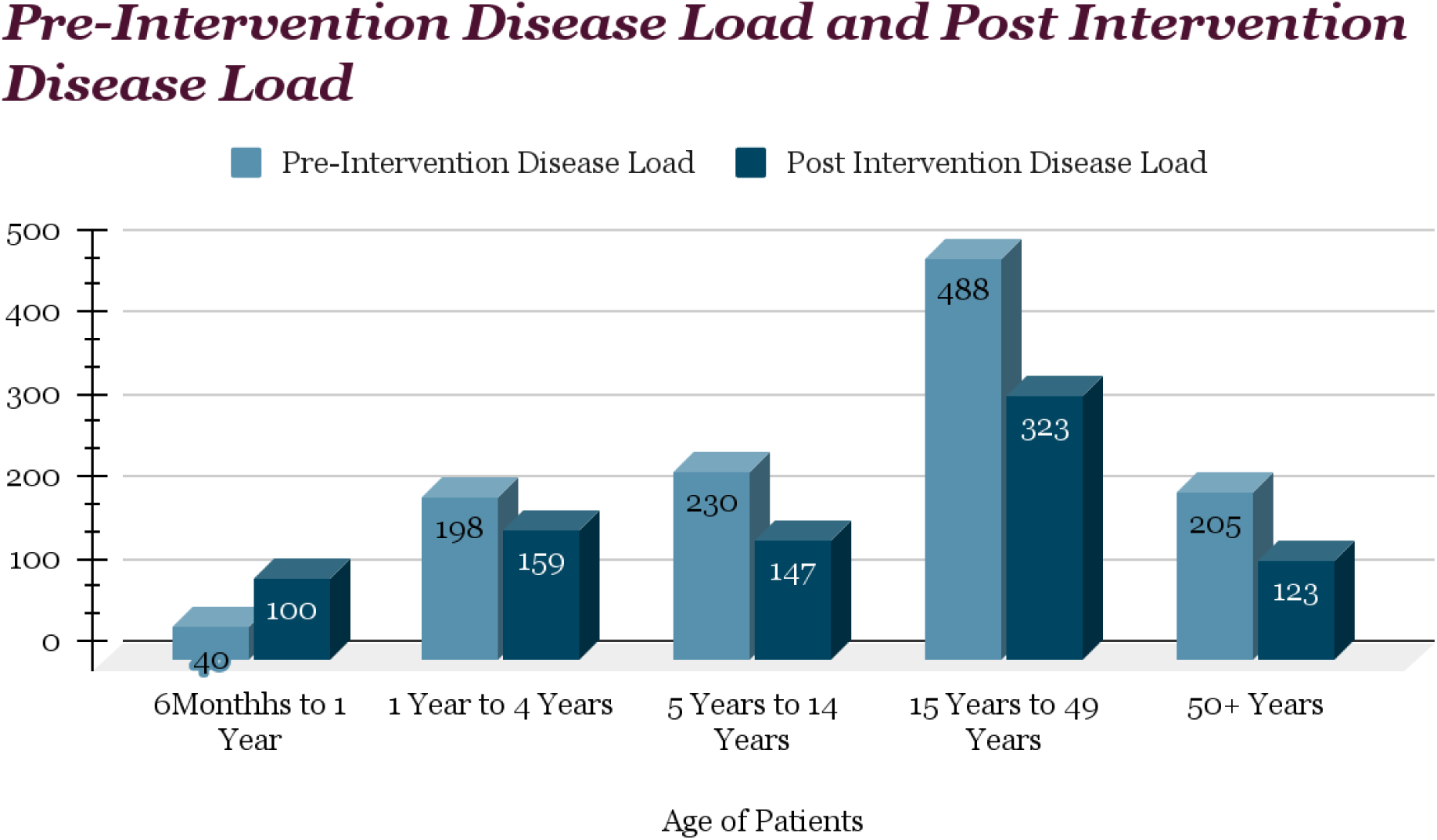
Age Based Disease Load Reduction.

Besides age-related interpretation of changing epidemiological trends, the gender analysis also deserves mention, because it is common assertion that women in rural areas are easy recipients of a disease keeping in view less education facilities and economic chores to improve quality of life. The biostatistics are,

No. of Male Patients Reported with RTIs in Pre Intervention phase= a1 = 587

No. of Male Patients Reported with RTIs in Post Intervention phase= b1 = 423

Decrease in No. of Male Patients Reported with RTIs= x1 = 587-423 = 164

Percentage Decrease=X1 = (x/a)100 = (164/587)100 = 27.93%

**Graph 4.3.**
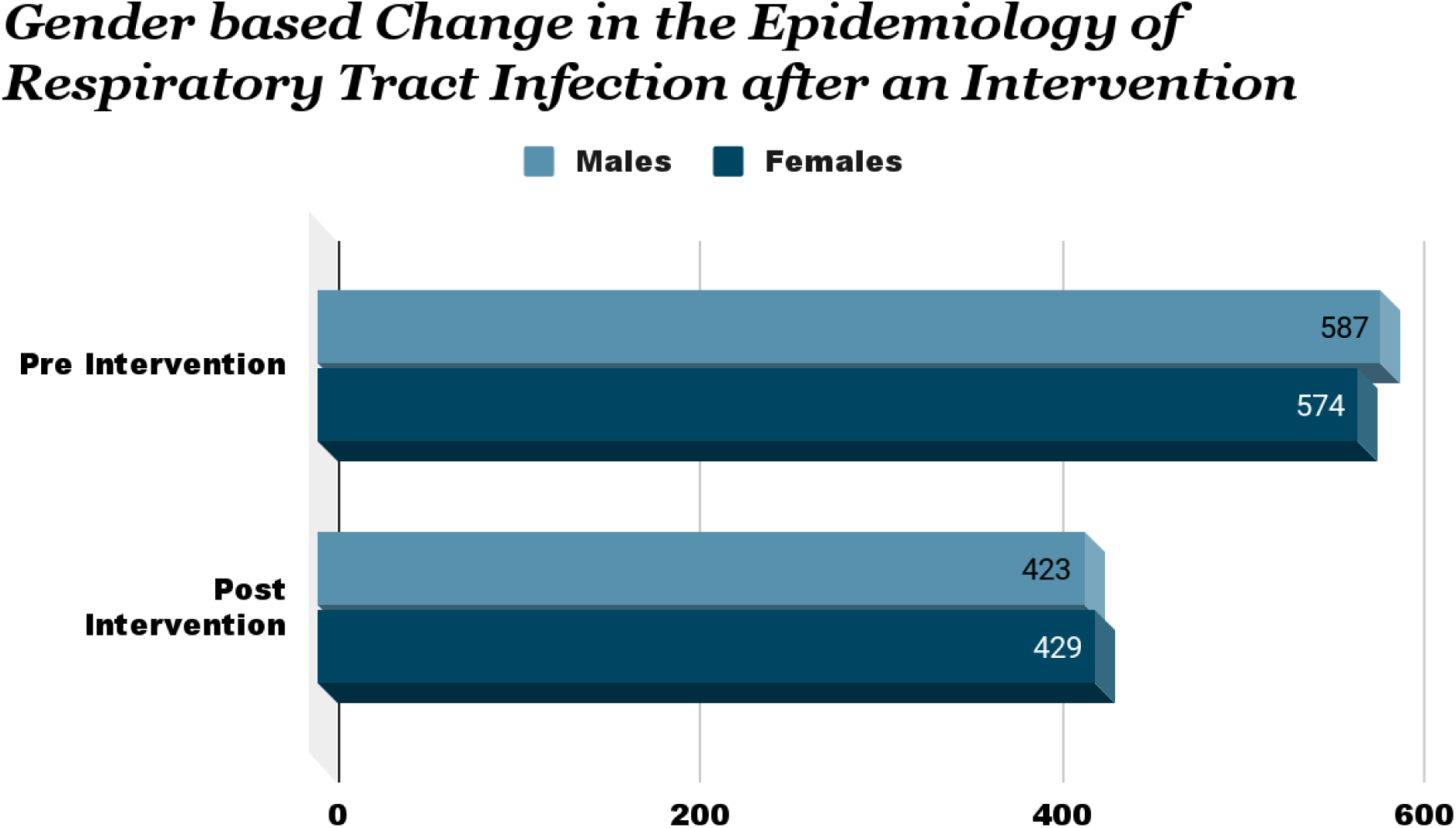
Age Based Disease Load Reduction.

No. of Female Patients Reported with RTIs in Pre Intervention phase= a2 = 574

No. of Female Patients Reported with RTIs in Post Intervention phase= b2 = 429

Decrease in No. of Female Patients Reported with RTIs= x2 = 574 - 429 = 145

Percentage Decrease= X2 = (x/a)100 = (145/574)100 = 25.26%

## Conclusion

Summing up, the project is conducted in a rural area comprising seven villages and provided by one basic health unit. The theoretical foundation of the study is intervention mapping while a mixed approach consisting of both the quantitative and qualitative is applied for data collection and program designing. Six subunits of intervention mapping are the philosophical constituents of the research while the whole scheme is undertaken in three phases. In the first phase of 45 days, data is collected to know the general occurrence of RTIs in terms of the number of patients reported in the outpatient department of the BHU. The commencement of comprehensive evidence based public health plan, developed after qualitative surveys in phase 1, entails the onset of phase 2.

This phase is the practical application of the plan coined with consultation after knowing the population acceptance. The public health project involves disease prevention strategies at individual and community level. The result portrays a general decrease in respiratory tract infections as almost 27% less number of patients are reported in the post intervention phase of the project. The mean trend remains the same in different age groups with some fluctuations. Moreover, the study also focuses on gender based biostatistics and a reduction in the burden of RTIs in both the genders is noticed. Furthermore, ethical consideration is given a prime weightage in conducting the research. This range of canvas paired with a unique research design elaborates the significance of the study.

## Data Availability

All data produced in the present work are contained in the manuscript

